# Testing mobile air purifiers in a school classroom: Reducing the airborne transmission risk for SARS-CoV-2

**DOI:** 10.1101/2020.10.02.20205633

**Authors:** J. Curtius, M. Granzin, J. Schrod

**Affiliations:** Institute for Atmospheric and Environmental Sciences, Goethe University Frankfurt am Main, Altenhöferallee 1, 60438 Frankfurt am Main, Germany

## Abstract

Airborne transmission of SARS-CoV-2 through virus-containing aerosol particles has been established as an important pathway for Covid-19 infection. Suitable measures to prevent such infections are imperative, especially in situations when a high number of persons convene in closed rooms. Here we tested the efficiency and practicability of operating four air purifiers equipped with HEPA filters in a high school classroom while regular classes were taking place. We monitored the aerosol number concentration for particles > 3 nm at two locations in the room, the aerosol size distribution in the range from 10 nm to 10 µm, PM_10_ and CO_2_ concentration. For comparison, we performed similar measurements in a neighboring classroom without purifiers. In times when classes were conducted with windows and door closed, the aerosol concentration was reduced by more than 90 % within less than 30 minutes when running the purifiers (air exchange rate 5.5 h^-1^). The reduction was homogeneous throughout the room and for all particle sizes. The measurements are supplemented by a calculation estimating the maximum concentration levels of virus-containing aerosol from a highly contagious person speaking in a closed room with and without air purifiers. Measurements and calculation demonstrate that air purifiers potentially represent a well-suited measure to reduce the risks of airborne transmission of SARS-CoV-2 substantially. Staying for two hours in a closed room with a highly infective person, we estimate that the inhaled dose is reduced by a factor of six when using air purifiers with a total air exchange rate of 5.7 h^-1^.

**Information Classification:** General

## 1. Introduction

Currently the transmission risks for viruses of the type SARS-CoV-2 pose a substantial challenge for all situations where people gather in closed rooms, be it in schools, shared offices, meeting rooms, restaurants, bars, etc. Especially in schools, where presence is obligatory and a high density of people in a room is frequently reached, a high level of responsibility rests with the authorities to ensure a safe environment for the school students and teachers.

Currently three transmission pathways are mainly considered important for Covid-19: direct transmission by droplets, airborne transmission by virus-containing aerosol particles and transmission via fomites (WHO, 2020). The direct infection by droplet transmission is well established as a transmission pathway for SARS-CoV-2 (WHO, 2020). Here it is assumed that large droplets (usually defined as droplets larger than 5 to 20 µm, WHO, 2020; Gralton et al., 2011) are transferred from one infected person to another, for example, by coughing, sneezing or speaking (see, e.g., Drossinos and Stilianakis, 2020; Vourinen et al., 2020). The droplets are assumed to be large enough to sediment to the ground quickly. Therefore, they do not travel more than 1.5 or 2 m horizontally. Prather et al., 2020, highlight the fact that also droplets much larger than 5-20 µm are able to stay airborne for many minutes and that the delineation between fast-settling droplets and aerosols should rather be made at 100 µm diameter. Morawska and Milton, 2020, recently compiled the evidence for airborne transmission. Their paper was accompanied by an open letter signed by 237 scientists supporting the view that the airborne transmission did not receive sufficient attention so far. The scientific evidence in favor of the airborne transmission pathway includes, for example, the description of infections that occurred over distances of many meters, such as several infections that occurred in a restaurant in China (Li et al., 2020), or 53 out of 61 attendees becoming infected during a choir rehearsal (Miller et al., 2020). Furthermore, aerosol samples containing viable SARS-CoV-2 were collected 4.8 m away from a Covid-19 patient in a hospital ward (Lednicky et al., 2020). Van Dormalen et al., 2020, showed that SARS-CoV-2 remains viable in aerosols with a half-life of 1.1 hours, and some virus remained viable for more than 3 hours. Despite of the mounting evidence in favor of airborne transmission an exact delineation of droplet and airborne transmission pathways is difficult (Tellier et al., 2019, Beggs, 2020; Jayaveera et al., 2020). For most infections it is not possible to reconstruct the details of the transmission. It is by now well-known that infected persons frequently stay completely asymptomatic or develop only mild symptoms that are not associated with a potential Covid-19 infection. Furthermore, infected persons are highly contagious shortly before symptoms of the disease occur (pre-symptomatic; WHO, 2020). Currently there are no well-established numbers to quantify the fraction of infections that are caused by the different pathways of infection (e.g., Jayaweera et al., 2020). These fractions are also expected to change depending on the effectiveness of measures and precautions that are introduced to reduce the transmission. Additionally, it is not known how many virus-containing aerosol particles have to be inhaled by a susceptible person to trigger an infection. Lelieveld et al. (2020) summarize the current knowledge, concluding that a dose of 100-1000 viral RNA copies have to be inhaled as the mean dose that causes an infection in 50% of susceptible subjects (D50).

People emit substantial amounts of aerosols and droplets when speaking, which could potentially contain the virus (Asadi et al., 2019; Asadi et al., 2020; Stadnytskyi et al., 2020). The amount of emitted aerosol particles increases with loudness. Furthermore, it was found that so-called super-emitters exist: 20% of the tested persons emitted far more particles than the average (8 out of 40 persons, Asadi et al., 2019). This led to the hypothesis that these super-emitters could also act as super-spreaders that are responsible for clusters with many infections occurring at single events (Asadi et al., 2020, Endo et al., 2020). The particles emitted when speaking have an average dry size of about 1 µm. It is not well known, in how far the particles emitted during speaking contain viruses if the speaker is infected with SARS-CoV-2. Lelieveld et al., 2020, argue that in case of highly infective emitters about 3% of the particles <5 µm contain virus RNA. But this number is currently not well-established. Stadnytskyi et al., 2020, report higher aerosol emissions from speaking than Asadi et al., 2019. When saying the words „stay healthy”, it was estimated that at least 1000 droplet cores that contain virions are emitted per minute with an average diameter of about 4 µm. The differences might be explained by the particularly high aerosol emissions when pronouncing the “th” in “stay healthy”. In schools and universities talking with a loud voice for longer periods during classes and lectures occurs frequently. Therefore, the aerosol transmission pathway could be of special importance. Longer periods of loud speaking by an infected asymptomatic or pre-symptomatic teacher, professor or student could lead to a high level of virus-containing aerosol particles in closed, unvented or insufficiently vented rooms even without coughing or sneezing. Simulations by Beggs, 2020, show that speaking in an office room can lead to similar levels of virus-containing aerosols as occasional coughing, and these levels could be high enough so that sufficient virus-containing particles are inhaled by other persons in the same room to cause an infection.

For a relative humidity far below 100 % droplets emitted during speaking will lose a large fraction of their water content rapidly and much smaller droplet cores will remain (e.g., Mikhailov et al., 2004). Droplet cores with sizes below about 10 µm will remain airborne for minutes to hours and these particles are transported in a room by thermal convection, turbulence and other air movements. Therefore, the particles are distributed throughout a classroom within minutes and they can accumulate in a closed room over hours. Mobile air purifiers offer the possibility to reduce the aerosol load in closed rooms substantially (Offermann et al., 1985; Shaughnessy and Sextro, 2006). If the air in a closed room is drawn continuously through a filter, the risk of an infection from respirable aerosols will likely be reduced (Offermann et al., 1985; Miller-Leiden et al., 1996). The Clean Air Delivery Rate (CADR) is used as a metric to characterize the efficacy of air purifiers (Shaughnessy and Sextro, 2006, Küpper 2019,). It is given as the product of the device’s particle removal efficiency η and the volumetric flow rate 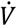 through the device. It can also be calculated by subtracting the natural particle decay rate *k*_*nat*_ in a room from the decay rate measured with the air purifier in operation *k*_*purifier*_ and multiplying by the volume of the room (Küppers et al., 2019):

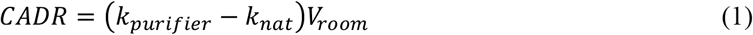

In order to clean the air as efficiently as possible from virus-containing aerosols, the CADR of the purifier should be as high as possible.

The experiments conducted in this study will test if air purifiers offer an efficient and realistic way to reduce the aerosol load during day-to-day operation in a high school classroom. The reduction of aerosol particles in general will also lead to a reduction of potentially virus-containing aerosols. The risk of infection through inhaled aerosol particles would therefore likely be reduced.

So far, only a few investigations on the operation of air purifiers under real life conditions exist. Küppers et al., 2019, study their performance in an office room environment. A few studies have tested air purifiers in classrooms, e.g. as a measure to reduce exposure to PM for children with asthma (Jhun et al., 2018),, the efficiency to remove ultrafine particles, black carbon, PM_2.5_ and PM_10_ was tested (Polidori et al., 2013), or PM_2.5_ and PM_10_ alone (Park et al., 2020).

Especially in a densely seated classroom it is necessary to take precautionary measures to prevent aerosol infections with SARS-CoV-2. This can be achieved by frequent venting by opening the windows or by filtration with air purifiers. Most schools in Germany are not equipped with pre-installed ventilation systems or Heating, Ventilation and Air Conditioning systems (HVAC). Venting is usually done by opening the windows during the breaks. By venting, the room air is diluted and exchanged with outside air within a few minutes. The exact exchange rates depend on numerous conditions such as wind speed and direction, temperature difference between outside and inside air, window size, etc. The venting reduces the infection risks considerably, but during winter time in Germany it is questionable whether it is feasible to vent every classroom several times per hour, each time for 5 to 10 minutes, as officially recommended by the German commission for indoor air hygiene (IRK, 2020a and 2020b). This would lead to uncomfortably cold classrooms, substantially increased heating costs and high CO_2_ emissions from the additional heating. Therefore, cleaning the air in addition to venting could be an important safety measure to reduce the concentration of potentially virus-laden aerosols in classrooms.

Note that we will not study the role of face masks as a measure to reduce the transmission of aerosols or droplets. For a discussion of this subject, see, e.g., Drewnick et al., 2020, or, Lelieveld et al., 2020. Furthermore, we will only study the role of air purifiers for the aerosol transmission pathway. It should be noted that in cases of a direct droplet or aerosol transfer between two persons in close proximity (’face-to-face’) the transmission risks cannot be substantially reduced by the operation of air purifiers that are located at some distance in the same room.

In this study we investigate if the operation of mobile air purifiers in classrooms can reduce the aerosol load fast, efficiently and homogeneously. A simple calculation is provided to estimate the average concentration of virus-containing aerosol in the closed room if an infected person is present that emits a high amount of virus-containing aerosol via speaking. Also the uptake of virus-containing aerosol via inhalation is estimated as a function of time. It is demonstrated that the uptake depends critically on whether or not air purifiers are operated in the room. Furthermore, we assess if the operation of air purifiers is hindered by other factors such as noise level or cleaning and maintenance of the purifiers.

## 2. Methods

This section describes the air purifiers that were used in our tests. Furthermore, the instrumentation for characterizing the aerosol is described, as well as the design of the tests that were conducted.

### 2.1 Air purifiers

The tests were performed with commercially available mobile air purifiers (Philips Model 2887/10), which are offered as regular household appliances. The air purifiers are equipped with HEPA filters (High Efficiency Particulate Air Filter) that remove more than 99.97% of the particles in the size range of 0.1 to 0.3 µm according to the manufacturer (DOE STD 3020 2015). Note that these filters are electret filters for which the filter efficiency decreases over time (Schumacher et al., 2017, Asbach et al., 2020). The volume flow through the purifier can be adjusted in five stages: ‘sleep’, 1, 2, 3 and ‘turbo’. According to the manufacturer the CADR is at least 333 m^3^/h when operated in ‘turbo’ mode and the device is designed for rooms of up to 79 m^2^. Note that the room size specified by the manufacturer is given with respect to the reduction of allergens and not with respect to SARS-CoV-2 reduction. Table 1 shows the measured volume flows for the higher flow regimes (stages 2, 3 or ‘turbo’) that were used during our tests. To reduce the risks of aerosol transmission, the volume flow through the purifier should be set as high as possible. Furthermore, we determined the energy consumption and the noise level during operation at these stages. The noise level was determined using two simple mobile phone apps (“Dezibel X” and “dB Meter”). We measured 1 m above the instrument but outside the main airflow exiting the instrument using the internal microphone of an iphone 8. The two apps agreed within ±0.5 dB. The air purifier’s dimensions are 24,0 cm x 35,9 cm x 55,8 cm.

**Table 1:** Technical properties of the air purifiers (Philips 2887/10) operated at a range of volume flows.

| ventilation flow stage | volume flow | noise level<br>(1 m above purifier) | power consumption |
| --- | --- | --- | --- |
| 2 | 186.7 m<sup>3</sup>/h | ~39 dB | 9.2 W |
| 3 | 256.6 m<sup>3</sup>/h | ~48 dB | 16.9 W |
| ‘turbo’ | 365.2 m<sup>3</sup>/h | ~54 dB | 42.8 W |

The purifier includes a simple pre-filter for coarse dust and aerosol (metal screen with mesh width ∼0,5 mm), as well as an active charcoal filter with screens with mesh width of ∼0,7 mm. Following the recommendation of the German commission for indoor hygiene we avoided air purifiers that rely on the use of ozone generators, ionizers, UV light, etc. (IRK, 2020a and 2020b).

When installing and positioning the air purifiers in the class room, several aspects should be considered. When just a single purifier is installed, the positioning should ideally be at a central place in the room (Kähler et al., 2020). If several purifiers are installed, the instruments should be distributed evenly in the room. Küpper et al., 2019, show that also a placement in the corners is possible as long as the flows towards the air intake and from the clean air outlet are not obstructed. Especially any blocking of free circulation, e.g. by placing the air purifier underneath a table, is reducing the efficiency of the purifier substantially. Obviously all safety aspects need to be considered, for example, emergency exits must not be blocked when installing the mobile air purifiers.

### 2.2 Instrumentation

#### Measurement of aerosol number concentration by condensation particle counters

For measuring the aerosol concentration several ultrafine Condensation Particle Counters (TSI, uCPC model 3776) were employed. The uCPCs allow a precise measurement over a wide range of particle concentrations (0.1 cm^-3^ to more than 100 000 particles cm^-3^). Ultrafine particles starting from a size of 2.5 nm are detected. The sampling was performed directly from the room air without using any additional sampling lines. The uCPCs use butanol as the working fluid for growing the aerosol particles to sizes at which they can be detected optically. The exhaust air of the uCPCs was transferred to the outside by use of an exhaust line. The sample flow was set at 1,5 l/min. The measurement uncertainty is about ±5%.

#### Measurement of aerosol size distribution and total aerosol mass (PM_10_)

The aerosol size distribution is measured over a wide range from 10 nm to 10 µm by applying a combination of a Scanning Mobility Particle Sizer (SMPS, consisting of an electrostatic classifier TSI model 3082 with Differential Mobility Analyzer TSI model 3080 and uCPC TSI model 3776) and an Optical Particle Sizer (OPS, TSI model 3330). The SMPS measures the aerosol size distribution in the size range 10 to 300 nm. The OPS measures the size distribution in the size range from 300 nm to 10 µm applying 16 size bins. The approximate total mass (PM_10_) is derived from the size measurements and assuming a mean particle density of 1.6 g/cm^3^ (Pitz et al., 2003).

#### CO_2_ -sensor

A CO_2_ sensor (NDIR) is used for monitoring the CO_2_ mixing ratio (Trotec model BZ30). The sensor measures the CO_2_ mixing ratios in the range from 0 to 10 000 ppm CO_2_. The uncertainty is given by the manufacturer as ±75 ppm. The instrument also records the temperature and relative humidity.

### 2.3 Measurement site and study design

A sketch of the placement of the air purifiers and the measurement instruments in the classroom is shown in Figure 1. The classroom (room B109 of Leibnizschule Wiesbaden) is located in the second floor. It has a length of 8.24 m, width of 6.18 m, a ceiling height of 3.66 m and a total volume of about 186.4 m^3^. Small additional recess volumes from the door and window area were not considered. Two rows of windows are located on one side of the room (Figure 1). Up to five windows of size 0.70 m x 1.36 m were fully opened for venting the room. In addition, the door (0.93 m x 1.99 m) was fully opened when venting. The room offers 27 places for the school students and a place for the teacher. For our tests, also a scientist was present to conduct and monitor the measurements. Three or four air purifiers were operated in the room simultaneously. For some of our measurements air purifier #2 (see Figure 1) was turned off. At the back side of the room the uCPC, SMPS, OPC and the CO_2_ -sensor were placed on a table. The second uCPC was operated on a desk located next to the teacher’s desk at the front side of the room. The air purifiers were placed directly on the floor and were distributed across the room as indicated in Figure 1.

**Figure 1:**
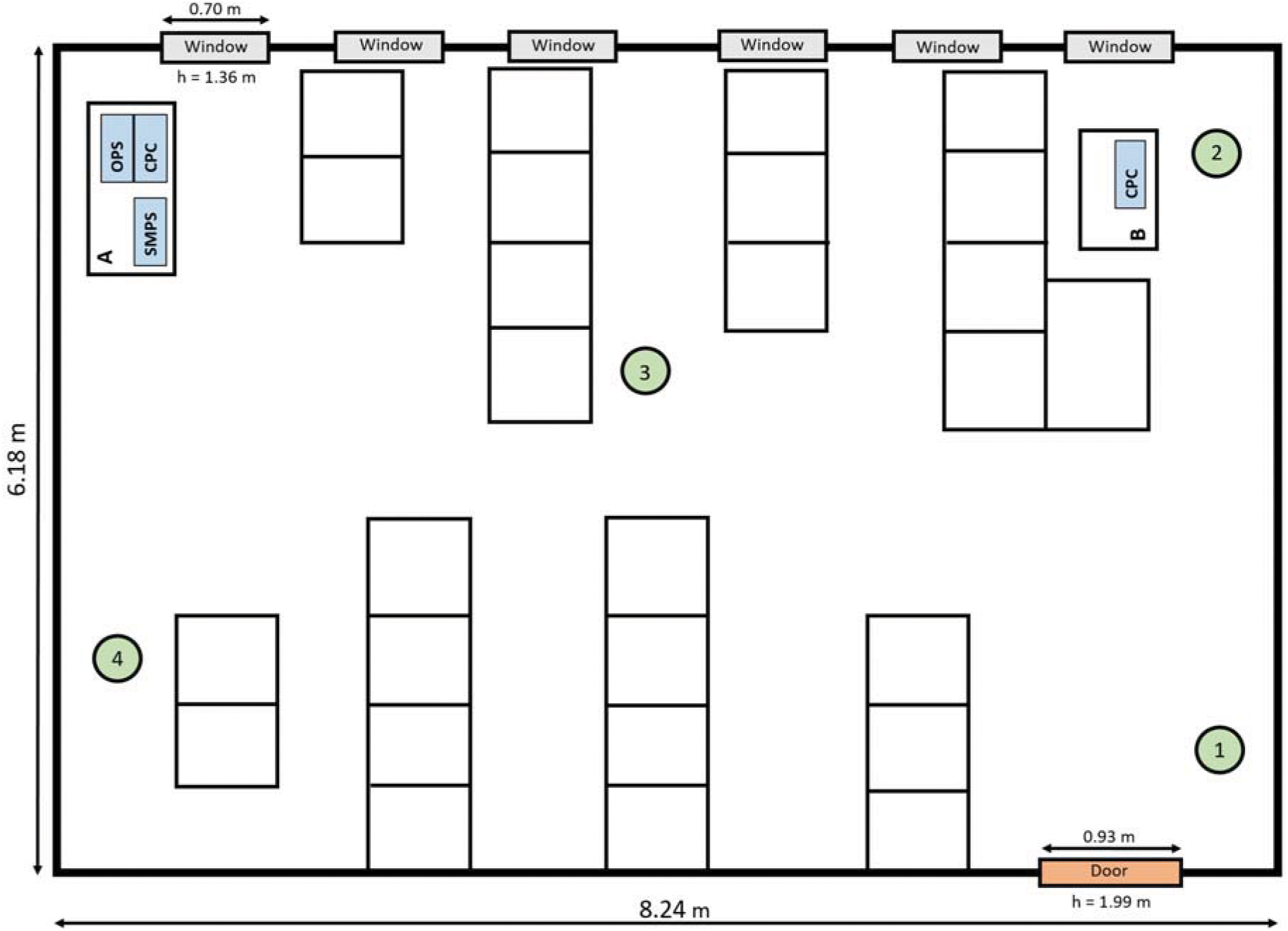
Sketch of the classroom indicating the position of the air purifiers (#1 to 4) and the measurement instrumentation at two locations A and B.

For comparison we also conducted measurements in a neighboring classroom (B110), but without operating any air purifiers. Here the air was monitored continuously with a uCPC and an OPS. The room dimensions were very comparable (8.25 m x 6.32 m x 3.69 m, total volume ∼192.4 m^3^). This room also has six windows of which up to five were fully opened for venting. Also in this class typically 27 students, one teacher and one scientist were present.

The two rooms are oriented perpendicular to each other, resulting in a difference between the rooms as the windows of the room without air purifiers were facing towards a busy road, while the windows of the other room opened towards a quiet side way of this road. Consequently, aerosol number and mass concentrations in the room facing the street were higher most of the time, even when purifiers were turned off in the other room.

The air purifiers were operated at the school from Monday, 14 September, until Friday, 18 September 2020. During this time 8 single lessons (45 min each) and two double lessons (90 min each) were held at the class room with the air purifiers and the measurement instrumentation running, while 18 lessons were held at the reference room, respectively. On Friday, only the purifiers were operated for 5 lessons without the aerosol instruments running in order to obtain a judgement on the noise levels produced by the purifiers alone without influence from the measurement instrumentation. Air purifier settings were varied by operating 3 purifiers at stage 3 on Monday and Tuesday, and then 4 purifiers at stage 3 for 4 lessons and at ‘turbo’ stage for 1 lesson.

### 2.4 Method for estimation of the concentration of virus-containing aerosol and inhaled dose

The decrease of the aerosol concentration in the classroom through filtering after particles enters from outside while venting the room by opening the window is not directly comparable to the situation of aerosol emissions by an infective person in the room. An infective person in a closed room that is continuously speaking acts like a continuous point source of aerosol particles containing virus RNA. In order to assess how the concentration of RNA-containing aerosol particles changes in a closed room with and without air purifiers, we performed a calculation with simplifying assumptions. We assumed an infected pre-symptomatic or asymptomatic person at a stage when he/she is highly contagious, e.g., in the hours before symptoms occur. This person was assumed to be frequently talking with a loud voice (50% of time, e.g., a teacher, lecturer or a student reading or presenting). In order to make our calculation comparable to a similar approximation that was recently published by Lelieveld et al. (2020), we based our calculation on the same assumptions. Note that several of these assumptions are currently highly uncertain for SARS-CoV-2 and may change in the future. Especially the amount of RNA-containing particles that form an infectious dose as well as the fraction of exhaled particles that contain virus RNA are uncertain.

Following the arguments of Lelieveld et al., 2020, we assumed an infectious dose D50 of 316 particles as the amount of virus RNA that has to be inhaled by a host to cause an infection at 50% probability. Furthermore, we assumed that aerosol particles are deposited in the respiratory system of the host at 50% probability. Breathing and speaking by an infective individual releases 0.06 and 0.6 particles cm^-3^, respectively, into the exhaled air (Asadi et al., 2019; Stadnytsky et al., 2020; Lelieveld et al., 2020). These assumptions describe rather the case of a super emitter (highly contagious, speaking a lot, emitting a high amount of particles with virus-RNA) compared to a more average case of an infected student that is not talking for a large fraction of the time. The speaking/breathing ratio is assumed to be 0.5 and the respiration rate per person is 10 liters per minute. We assumed the concentration of virus RNA present in the exhaled particles to be 5 × 10^8^ per ml for a ‘highly infective’ person (Lelieveld et al., 2020). The particles are exhaled with an average wet diameter of 5 µm when exiting the mouth, but due to the rapid evaporation at lower humidity the particle size quickly reduces to 1-2 µm. The 1/e-lifetime for viable SARS-CoV-2 in aerosol is assumed to be 1.7 hours (van Dormalen et al., 2020). With these assumptions Lelieveld et al., 2020, derived an emission of 198 000 particles per hour from the emitting person and about 3% of the particles emitted during breathing and speaking contain virus RNA. For a classroom of 180 m^3^ volume with a low air exchange rate of 0.35 h^-1^ and without face masks being used, their model predicts a steady-state concentration of RNA-containing aerosol of 0.034 l^-1^. For the highly infective case, a susceptible person in the room for 2 hours will take up a dose of 22 RNA-containing particles, representing a 4.7% risk of becoming infected. In a room with 25 persons, this leads to an overall risk of 70% that at least one of the other 24 persons becomes infected. For an air exchange rate of 5.7 h^-1^ the risk per person reduces to 0.7 % and 16.5 % that at least one of the persons in the room becomes infected. All the details of the assumptions, parameters and equations used are discussed in Lelieveld et al., 2020.

We performed a similar calculation as derived by Lelieveld et al., 2020, as we assumed the situation of a classroom for two hours without any ventilation and compared it to a situation with air purifiers running (air exchange rate 5.7 h^-1^). Similar to Lelieveld et al., 2020, we did not perform a detailed flow calculation of the air movements, turbulent mixing and dilution processes, etc. Instead, we assume an instantaneous homogeneous mixing of the emitted aerosol in the room, which seems to be a justifiable assumption based on the regularly almost identical concentration measurements of the two uCPCs located at different positions in the room (see Results and Discussions below and Figure S1). For the typical classroom we assumed a volume of 180 m^3^ and a volume flow rate of the four air purifiers totaling 1026 m^3^/h and for simplicity assuming a 100% filter efficiency (CADR = volume flow rate). For comparison we also performed calculations assuming that the room is vented by opening the windows every 20 minutes (Fig. S2 and S3).

To confirm the validity of our estimation experimentally we performed a set of test measurements in a seminar room (128 m^3^) without people. Here we placed an aerosol generator in the middle of the room, producing a constant output of NaCl solution droplets. We then ran two experiments monitoring the number concentration of particles >3 nm with 3 uCPCs at different locations in the room and measuring the particle concentration >300 nm with 2 OPS instruments. In the first experiment all windows and the door were closed, we removed the pre-existing particles in the room by operating several air purifiers down to a total level <100 particles per cm^-3^ and then we switched off the purifiers and started the aerosol generator remotely. We observed the steady increase of particle concentration in the room over a period of 50 minutes. In the second case we continued to operate 3 purifiers when switching on the aerosol generator.

## 3. Results and Discussion

### 3.1 Aerosol measurements and air purifier performance

Figure 2 shows a representative measurement of the total aerosol number concentration (uCPC), the number concentration of large particles (0.3 to 10 µm, OPS), and the total aerosol mass (PM_10_, OPS) for the two rooms during a school lesson with windows and doors closed. The total number concentration in the room without purifiers decreased slowly over time and was reduced by about 30% at 12:06 when a window was opened and additional particles entered the room from the outside. The decrease in particle concentration while the room was closed was mainly caused by diffusion of the particles to the surfaces in the room, as well as coagulation processes and sedimentation losses. A fraction of this decrease was also caused by the respiration of the 29 persons in the room as a fraction of the aerosol particles was inhaled and deposited in the upper and lower respiratory tracts of the people in the room. The overall reduction was also found to vary from lesson to lesson (with the time constant of the exponential decay varying between 47 and 71 minutes) mainly because the loss processes are dependent on the particle size, the humidity in the room, the charging state of the aerosol, the presence of electrostatically charged surfaces in the room, and other factors.

**Figure 2:**
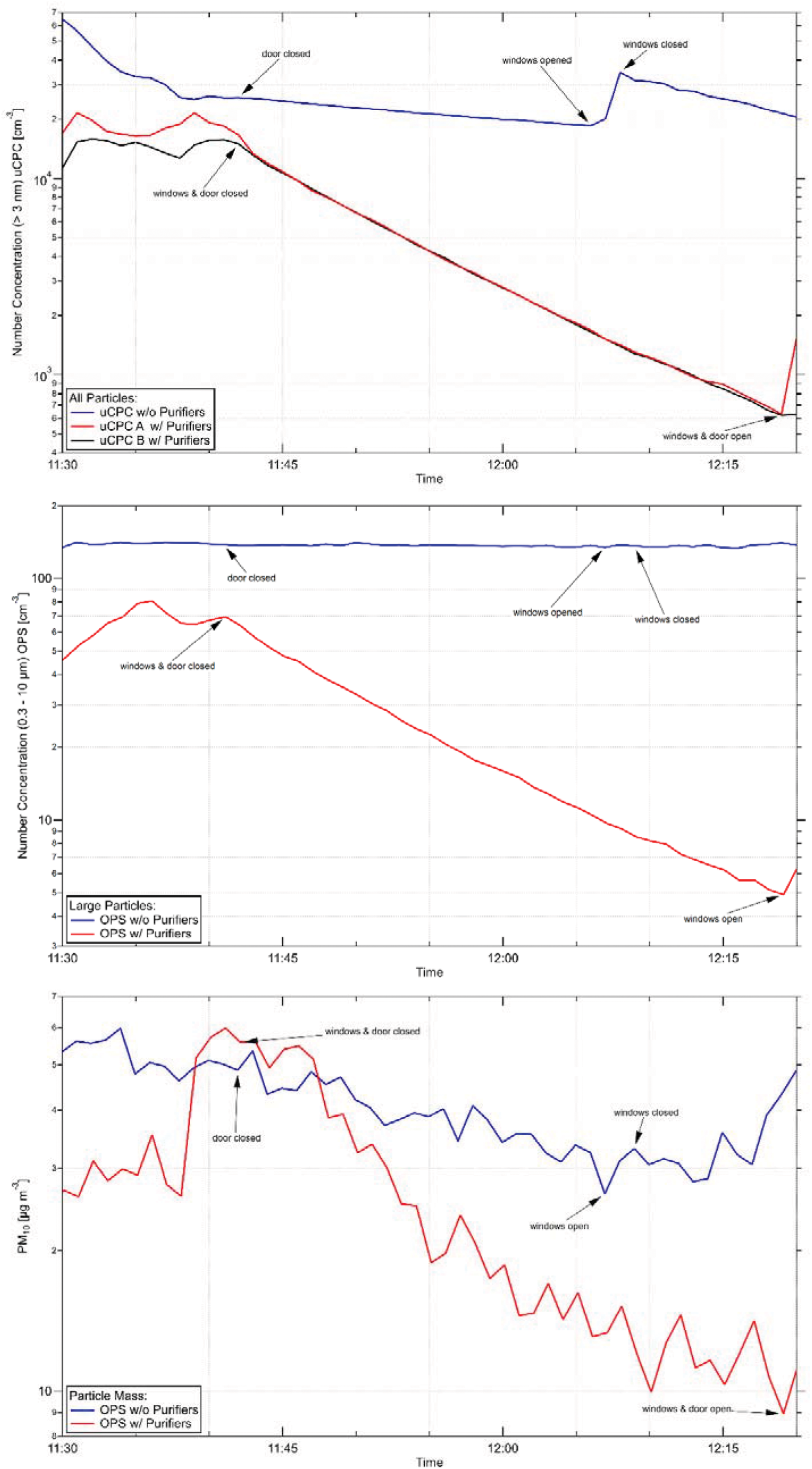
Top panel: Representative measurement of the total aerosol number concentration in the classroom with air purifiers (red and black line) and in the room without purifiers (blue line) during a lesson. Four air purifiers were operated at stage 3 during a lesson with windows and doors closed. A window was briefly opened in the room without purifiers at 12:06 for ∼1 minute, when additional particles flowed in from outside. Particle concentrations are averaged over 1 minute intervals. Middle panel: Number concentration of larger particles (0.3 to 10 µm, OPS measurement) in the classrooms with (red) and without (blue) air purifiers. Bottom panel: The particle mass concentration PM_10_ in the rooms with and without air purifiers. Values are more scattered due to low counting statistics for the largest particles that contribute most to the derived mass concentration.

The aerosol concentration decreased considerably more in the room with the air purifiers. The four purifiers were operated at stage 3 yielding a total volume flow of 1026.4 m^3^/h and an air exchange rate of 5.5 h^-1^. The aerosol concentration decreased by more than 95% within 37 minutes following an exponential decay rate. Both uCPCs measured almost identical values (red and black line in Figure 2a). This shows that the room was well mixed and the reduction of particles in the room was very homogeneous. This was very reproducible and we did not notice any parts of the room to be excluded from the action of the four purifiers. From the average of the three lessons where the four purifiers were operated at stage 3, the exponential decay of the total aerosol concentration as measured by the uCPCs, the decay constant is derived as 0.107 ± 0.01 min^-1^. Similarly from the decays in the other class room without purifiers an average value for the natural decay constant without air purifiers is determined as 0.020 ± 0.01 min^-1^. By using equation (1) the experimental CADR is derived for the four purifiers as 973 ± 150 m^3^/h. This is about 5% lower than the calculated CADR of 1025.3 m^3^/h from the measured volume flow of 4 × 256.4 m^3^/h and the stated filter efficiency of 99.97% but within the uncertainty of our measurements.

The total number concentration as well as the total mass from the OPS measurements in the two rooms are shown in the lower panels of Figure 2. The number concentration of particles in the range 0.3 to 10 µm decreased exponentially with a similar time constant as the total number concentration measured by the uCPC. In the room without purifiers the number of particles in the range 0.3 to 10 µm remained almost constant.

The total aerosol mass was reduced from about 35 µg/m^3^ at the beginning of the lesson to about 6 µg/m^3^ after about 37 minutes, while the total mass reduced to 20-25 µg/m^3^ in the room without purifiers. Note that a window was opened for about 1 minute at 12:06, leading to an increase in particle mass and total particle number concentration. The particle mass and particle number of larger particles measured with the OPS was systematically higher in the room without purifiers, as explained above. The OPS total number concentration for the reference room (blue line, Figure 2, middle panel) did not show a clear reduction like the uCPC data for this lesson (blue line, upper panel) and the OPS mass data (blue line, lower panel). This is caused by the fact that the OPS total number concentration is dominated by the smallest size channel which stayed constant over this lesson, while a slight reduction was seen in the largest size-channels, which have a dominating influence on the total mass concentration. Nevertheless, we cannot offer a full explanation why the smallest size channel of the OPS stayed constant during this particular measurement.

Figure 3 shows a composite plot of uCPC measurements from various school lessons in a closed classroom. All measurements were normalized to a starting level of 10 000 particles cm^-3^. The blue line shows a typical slow decrease when no purifiers were used. The decays of the total particle number concentration for the tests with air purifiers were highly reproducible for the different air exchange rates with 3 and 4 air purifiers. A halving of the particle concentration was reached in 10.0, 7.0 or 5.4 minutes (green, black and red lines, respectively), depending on the total flow of the purifiers.

**Figure 3:**
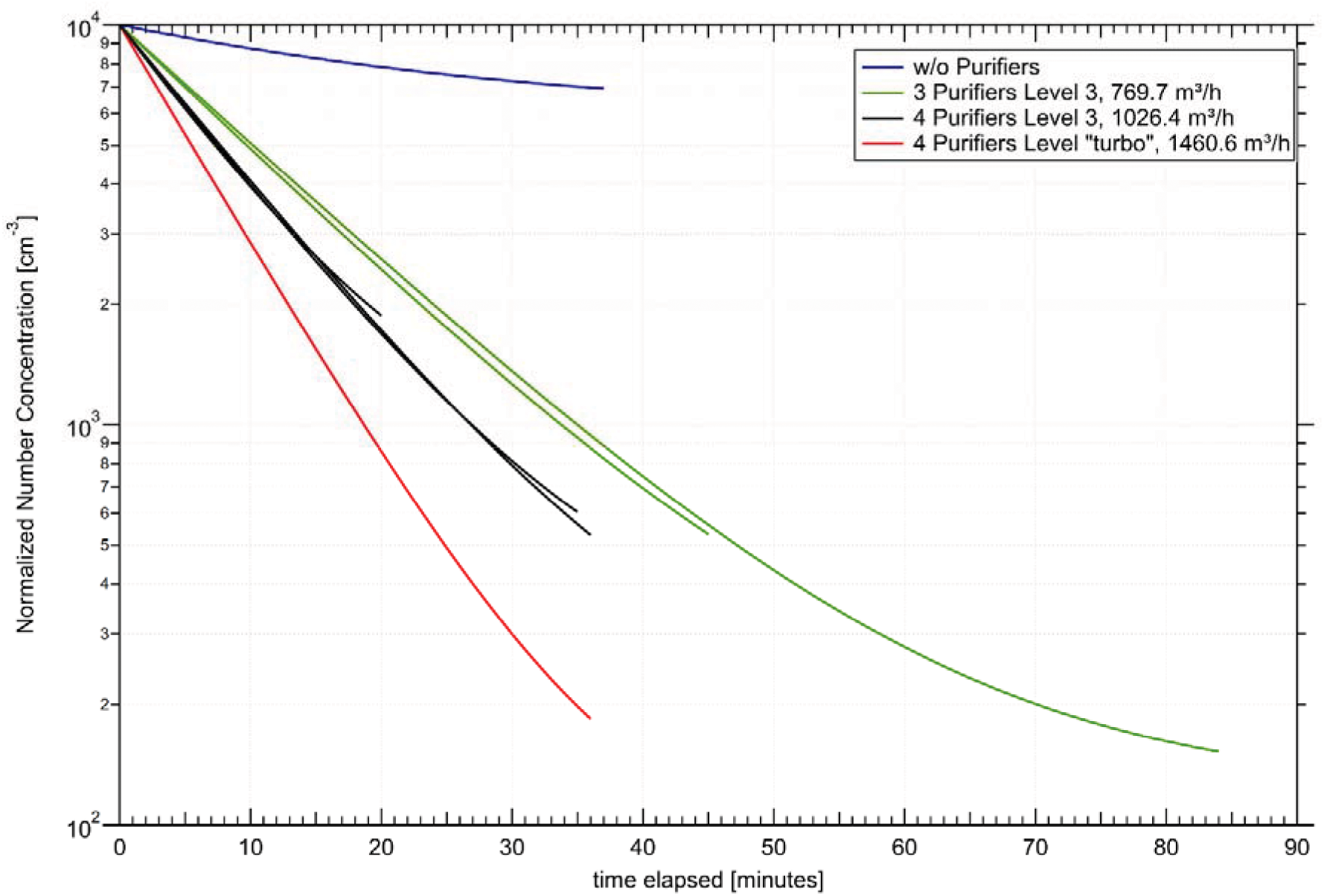
Reduction in aerosol particle concentration in a closed classroom without air purifiers (blue line) and with 3 or 4 air purifiers operating at stage 3 (3 × 257 m^3^/h per purifier, green lines; 4 × 257 m^3^/h per purifier, black lines) or stage ‘turbo’ (4 × 365 m^3^/h, red line). Data are normalized to a starting value of 10 000 particles cm^-3^. Data are displayed for the time intervals until door or windows were opened again.

Figure 4 shows the measurements of the SMPS instrument for particle sizes between 10 and 300 nm. The concentration levels (indicated by the color coding) decreased markedly for all sizes over time. Similarly, for all size bins measured with the OPS, the size resolved particle concentrations were decreasing evenly over time (Figure 5). The homogeneous reduction with respect to all particle sizes was confirmed by the fact that the mean particle size stayed constant at a value of ∼0.4 µm (pink dashed line).

**Figure 4:**
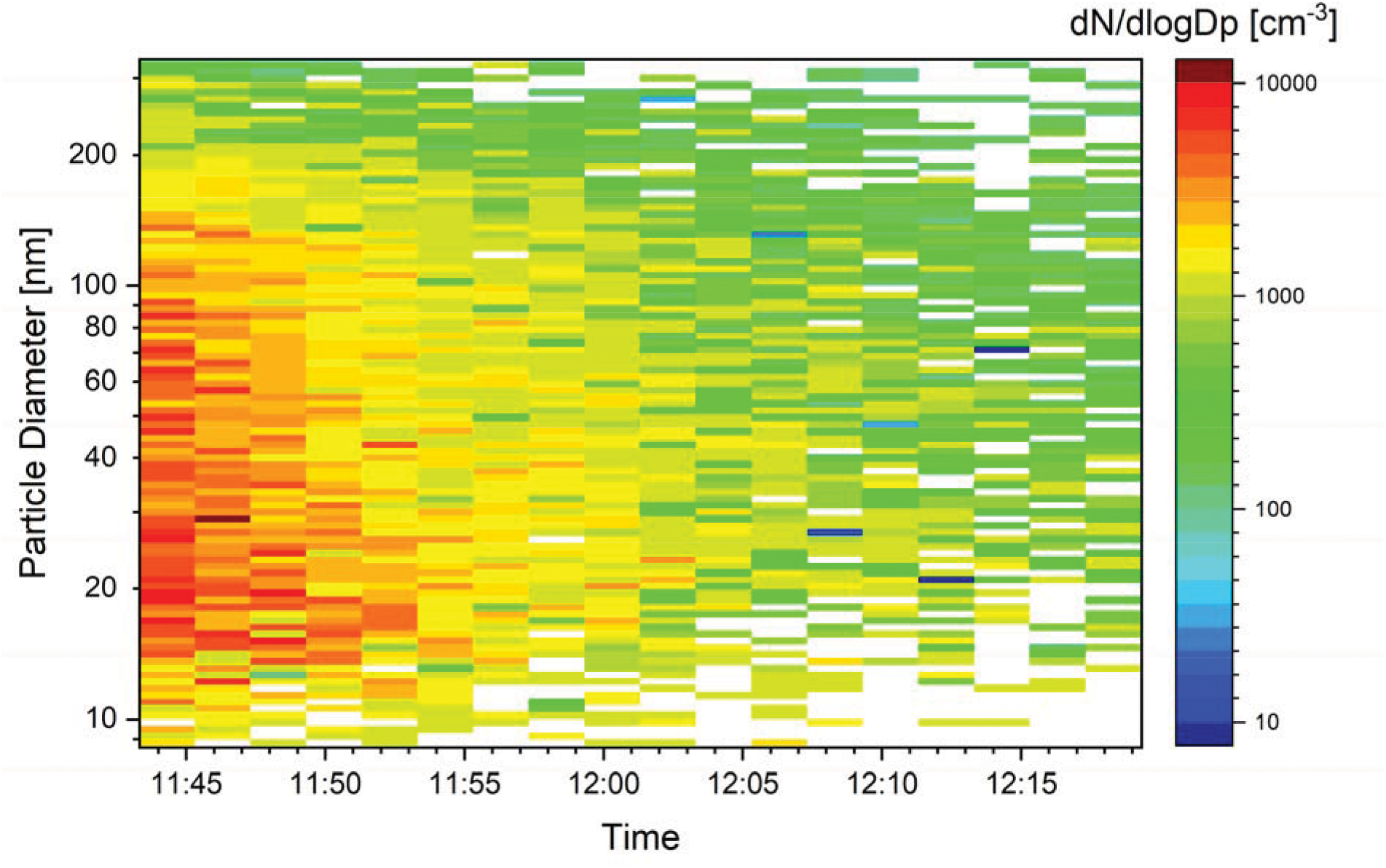
Measurement of the particle size distribution in the size range 10-300 nm as a function of time in the room with air purifiers (same lesson as in Figure 2). Red and yellow colors indicate high concentrations while green and blue colors indicate low concentrations. Particles <300 nm are filtered effectively and homogeneously.

**Figure 5:**
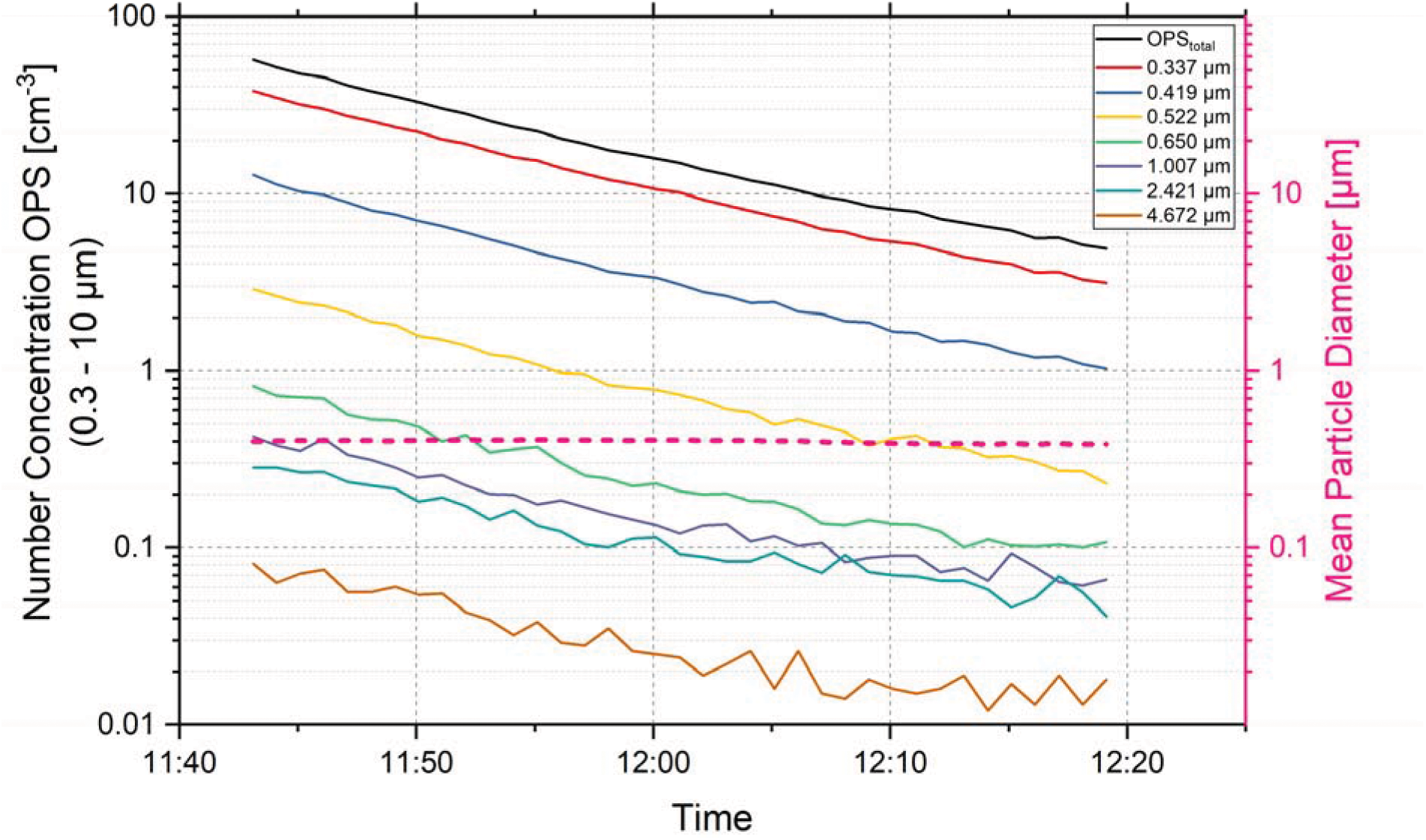
Total (black line) and size resolved decrease of particle concentrations for seven aerosol size bins of the OPS in the size range 0.3 to 5 µm (same lesson as in Figure 2). The mean particle diameter (dashed pink line) remains constant, indicating that all sizes decrease at the same rate.

Overall it can be stated that the use of air purifiers with HEPA filters decreased the aerosol load strongly within the time intervals between venting by opening the windows. The homogeneous reduction of all particle sizes indicates that, in case of an infectious person being present in the room, also the virus-containing aerosol particles emitted by this person from speaking or breathing will be reduced in the room air.

The concentration of virus-RNA containing aerosol particles for the case of a highly infective person (same assumptions as Lelieveld et al., 2020) is shown in Figure 6. We compare the situation with and without operating the air purifiers. It can be seen that a steady state concentration of about 0.006 particles per liter is quickly reached when the air purifiers are switched on, while without purifiers the concentration increases steadily reaching 0.069 l^-1^ after 2 hours. The inhaled dose for a susceptible person in the room increases over time. It reaches a value of 21 virus RNA units after 2 hours. With the purifiers running, the inhaled dose is 3.3 particles after 2 hours. These results are very comparable to the calculations obtained from the model by Lelieveld et al., 2020. As our results for the concentration levels and inhaled dose are within about ±10% of the results calculated with the model of Lelieveld et al., we assume that the infection risks stated above can also be applied for our results.

**Figure 6:**
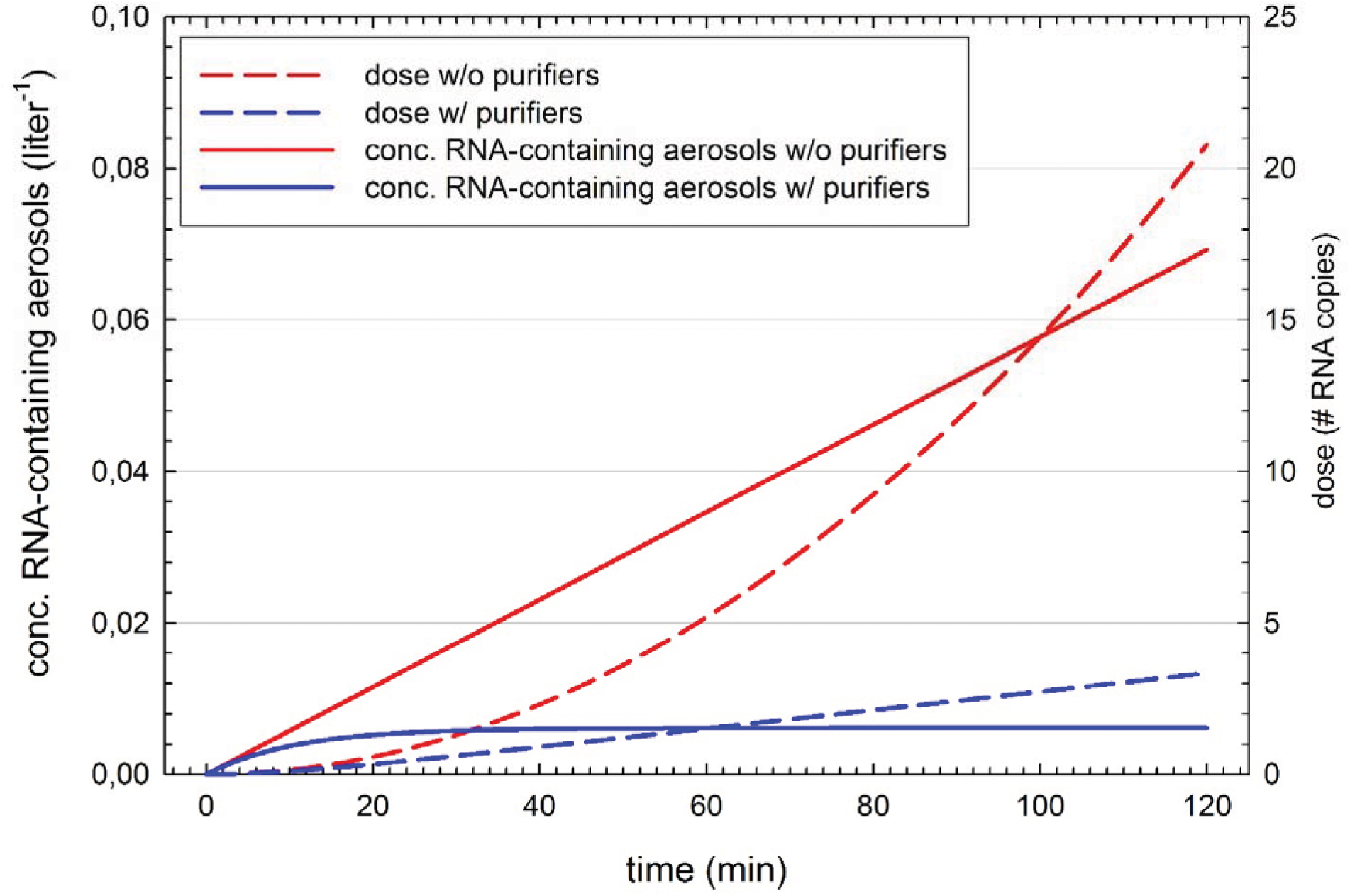
Estimated concentration of aerosol particles containing virus-RNA in a closed classroom (180 m^3^), in which we assume that a highly infective person emits on average 0.6 particles cm^-3^ of exhaled breath through loud speaking 50% of time and 0.06 cm^-3^ by breathing (red line without purifiers, blue line with purifiers) with an air exchange rate of 5.7 h^-1^. The dashed lines show estimates of the inhaled dose of virus-RNA units that is taken up by a person in the same room for two hours.

After 2 hours, the concentration of aerosol particles containing virus RNA in the room is more than 10 times higher ‘without purifiers’ compared to ‘with purifiers’. The difference between the two cases increases over time and it becomes larger if higher ventilation rates of the purifiers are realized. Similarly, the difference between the inhaled dose increases over time. After one hour the difference between the cases with and without purifiers is a factor of 3.5 and it becomes a factor of 6.3 after 2 hours. Note that the steady-state concentration does not depend on the room size. It is just given by the ratio of the particle source rate over the removal rate. A comparison of the effect of frequent venting by opening windows every 20 minutes to the use of air purifiers is shown in the Supplemental Online Material. If a highly efficient venting is achieved and reducing thereby 99% of the exhaled aerosols, the venting is about as efficient as the use of air purifiers. Ideally both measures are combined, yielding a dose after 1 hour that is 82% lower than in the case of a closed room.

This estimate is thought to illustrate the profound differences in a room without ventilation vs. a room equipped with purifiers with HEPA filters. Although several of the numbers used for this estimation are uncertain, we expect the finding to be robust that the difference in concentration levels and inhaled dose between ‘with’ and ‘without’ purifiers increases over time. The longer other susceptible persons are in a closed room together with an infective person, the higher the risks of airborne transmission even if the persons are separated by more than 2 m distance.

The results from our measurements in a seminar room to test the validity of our estimates of the development of the aerosol concentration are shown in Figure 7. When assuming a constant emission from the aerosol generator of 1.8 million particles per second, then the measured concentrations in the room are well described by the simple model for both cases, with and without the purifiers running. Especially in the case without purifiers the circulation and mixing in the room is not as strong and the uCPC measurements show differences in the concentration levels, indicating that the room air is not well-mixed throughout. Nevertheless, all three uCPCs measure the strong increase over time and the model describes the increase well. Although the emissions by the aerosol generator are orders of magnitude higher than the emissions by a person from speaking, these experiments show that the basic assumptions of a nearly well-mixed room (see e.g., Shaughnessy and Sextro, 2006) are approximately correct, even in a case when the room is not actively ventilated.

**Figure 7:**
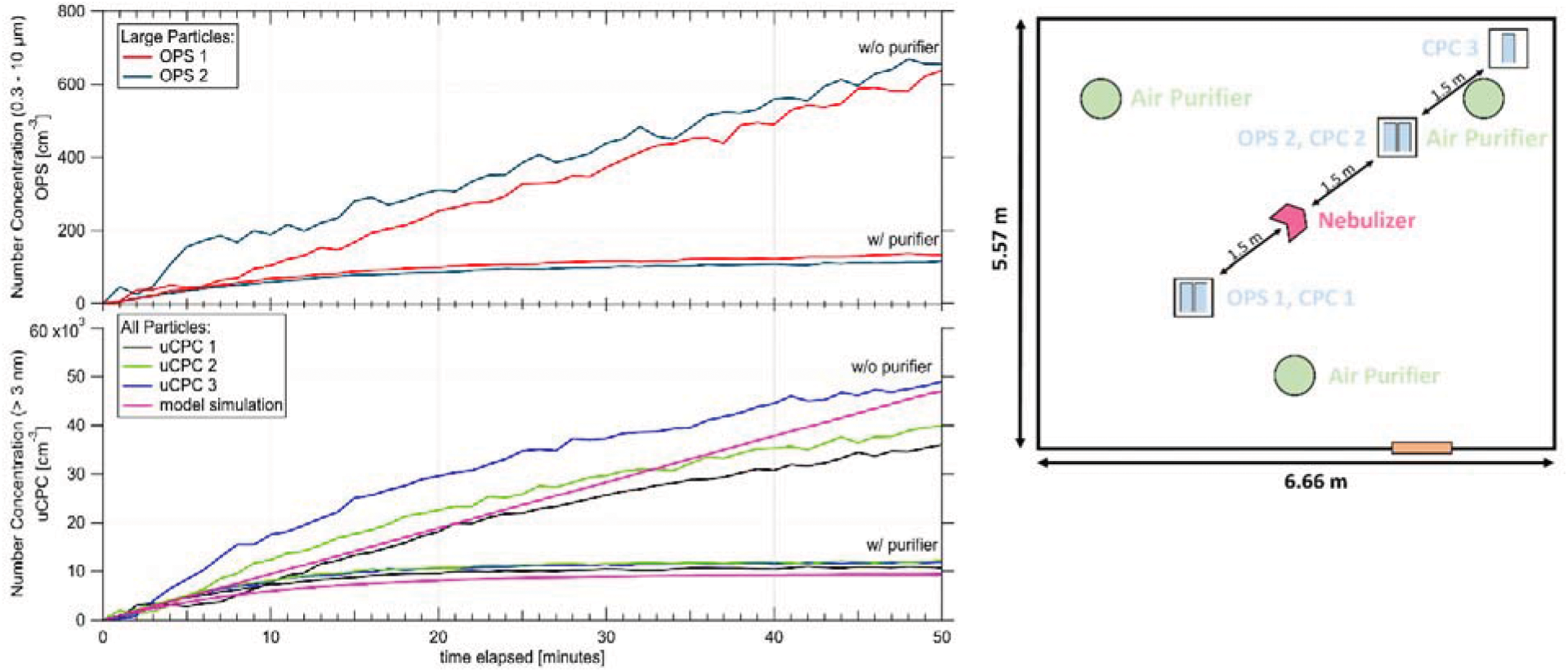
Left panels: Measurement of particle increase in a closed room without people, with and without purifiers. An atomizer is operated as a continuous particle source of NaCl solution droplets. 3 uCPCs and 2 OPS instruments measured at different positions in the room. Assuming an emission rate of 1.8 million particles s^-1^ a good agreement is reached between the model calculations and the measurements. Right panel: Sketch of the position of the aerosol generator (red), the three uCPC and OPS instruments (blue) and the position of the air purifiers (green) in the seminar room.

### 3.2 Carbon dioxide

Typical CO_2_ mixing ratios as measured during a school lesson with closed windows and doors are shown in Figure 8. The CO_2_ mixing ratio increased by 48 ppm per minute. At the end of the lesson a value above 2700 ppm was reached. It was repeatedly observed that even after several minutes of venting the room with open windows, the CO_2_ concentration did not fall below 1000 ppm. Therefore, already at the beginning of the lesson, the mixing ratio in the room was around 1000 ppm. The current recommendation by the German Environment Agency is that rooms should be vented at concentrations above 1000 ppm and have to be vented at values above 2000 ppm, as such high values cause headache and tiredness (UBA, 2009). For classrooms with a high density of persons this implies that also during lessons venting has to take place. This is independent of the use of air purifiers.

**Figure 8:**
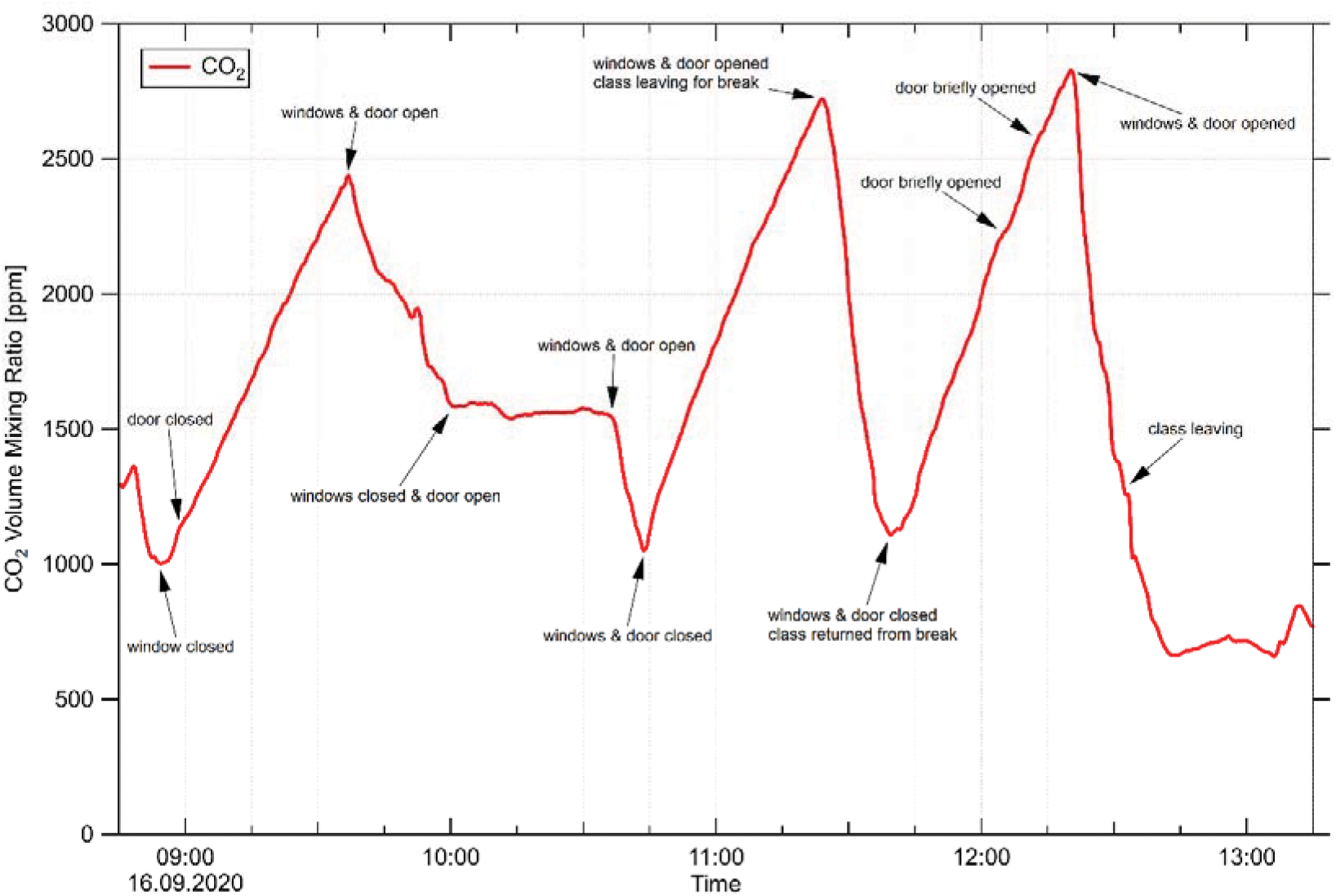
CO_2_ mixing ratio as measured in class during a school day. Even after venting the room for several minutes with door and windows wide open, CO_2_ levels do not drop below 1000 ppm. With classes proceeding in the closed room, CO_2_ levels quickly rise to mixing ratios of 2500 to 2800 ppm at the end of the lesson.

### 3.3 Noise levels

Besides the simple measurement of the noise levels (Table 1) we also conducted a survey among the school students and teachers. Care had to be taken that the noise level from the aerosol measurement instruments did not influence the impression of the noise level from the air purifiers. Therefore, we conducted some lessons with the purifiers switched on but without aerosol measurements. During the 5 lessons before the survey was conducted four purifiers were operated at stage 3. The students (age 14 to 15) did not consider the noise level as disturbing. From the 26 students that participated in the survey, 58% felt “not disturbed at all”, 27% felt “not disturbed” and 15% were neutral with respect to the noise level. From the 6 teachers that participated in the survey, 1 felt “strongly disturbed”, 2 felt “somewhat disturbed”, 1 was neutral, 1 felt “not disturbed” and 1 was “not disturbed at all” (see Figure S4). Only a very small number of teachers was included in the survey, so these numbers might not be significant, but the noise produced by the purifiers at the necessary high ventilation rates needs to be considered carefully before the procurement of purifiers. It should also be noted that the individual level of disturbance may change with the length of exposure time and it is probably dependent on the distance of the individual seating places from the purifier.

### 3.4 Cold drafts

None of our surveys revealed that the students or teachers were disturbed by cold drafts or the enhanced air circulation in the room. Here it should be noted that the tests were conducted during a phase of relatively high outside temperatures of 22 to 29 °C. As the purifiers blow the filtered air directly upwards, also persons that are seated close to a purifier are usually not affected by strong drafts.

### 3.5 Cleaning and maintenance

When running the air purifiers for a longer time (e.g., several hours of daily operation for several months during winter) a proper and regular cleaning and maintenance needs to be included. A visual inspection of the filters at the end of the week of operation in the classroom showed that the pre-filter and the active charcoal filter already had aggregated substantial amounts of coarse dust (see Figure S2). No deposits were visible on the HEPA filter from inspection with the naked eye. Many of the commercially sold air purifiers issue a warning signal when the filters need to be cleaned or exchanged. Nevertheless, safety precautions need to be observed when filters are cleaned or changed as the aerosol deposited on the filters may still contain infectious aerosol. Here it seems advisable that the filters are cleaned at a different place. If the HEPA filters are protected from the coarse dust (>10 µm), the HEPA filters should be operational for many months. Note that the commercial purifiers tested here did not contain proper pre-filters to fully retain the coarse dust particles >10 µm. Ideally two additional pre-filter stages with filter classes F7 and F9 should be included in a proper purifier system.

### 3.6 Co-Benefits

Our measurements as well as previous studies (e.g., Polidori et al., 2013; Chen et al., 2015; Park et al., 2020) demonstrate that operating air purifiers continuously in a closed room also reduces the amount of particulate matter (PM_2.5_ and PM_10_) considerably. The WHO recommends that the average exposure levels to PM_2.5_ should be below 10 µg/m^3^ because higher exposure increases the risks of ischaemic heart disease, chronic obstructive pulmonary disease, lung cancer, cerebrovascular disease leading to stroke, and various other diseases. Long-term exposure to high levels of PM_2.5_ reduces the life expectancy considerably and such high PM levels are among the leading health risk factors in many parts of the world (Lelieveld et al., 2019). Therefore, the average PM_2.5_ levels that students and teachers are exposed to should be kept below 10 µg/m^3^. Installing air purifiers would greatly help to reduce the average exposure to PM_2.5_. Similarly, exposure levels to various airborne allergens would be reduced (e.g., Park et al., 2017).

## 4. Summary and Conclusions

Air purifiers can reduce the aerosol load in a classroom in a fast, efficient and homogeneous way. In situations when windows and doors are closed for a longer period of time a large reduction in the inhaled dose of particles containing virus RNA is achieved and therefore the risk of aerosol infection is likely to be lowered. Staying for two hours in a closed room together with a highly infective person, we estimate that the inhaled dose via airborne transmission is reduced by a factor of six when using air purifiers with an air exchange rate of 5.7 h^-1^. The air purifiers should be equipped with HEPA filters (DOE STD 3020 2015, H13 or H14) and a high CADR of around 1000 m^3^/h or higher should be applied. In order to achieve high air exchange rates and homogeneous mixing in the entire room it can be of advantage to install several smaller purifier units. In addition to the HEPA filters, the purifiers need to be equipped with pre-filters to remove the coarse dust efficiently and the pre-filters need to be cleaned or exchanged regularly. If applied in school rooms, the noise levels from operating the air cleaners need to be considered. While large ventilation rates are desirable, the noise level needs to be sufficiently low in order to not disturb the ongoing classes.

In summary, the operation of mobile air purifiers in classrooms seems feasible as a practical measure that can quickly be implemented during an epidemic. In order to reduce the risks of aerosol transmission for SARS-CoV-2 air purifiers can form an important additional measure of precaution, especially in cases where no fixed ventilation systems are installed and when windows cannot be opened properly. The implementation and maintenance costs need to be compared to the substantial advantages of reducing the amount of infections and Covid-19 cases, the reduced needs for contact tracing and the avoidance of major disruptions caused by school closures. Nevertheless, air purifiers do not replace other measures for the reduction of transmission such as wearing face masks, hygiene measures and social distancing. The purifiers should be considered as efficient additional measures. An important co-benefit of a standard operation of air purifiers is that average levels of particulate matter (PM) are considerably reduced leading also to a long-term health benefit.

Rooms with a high density of people require frequent ventilation to reduce the CO_2_ mixing ratio. CO_2_ monitors should be used in order to ensure that CO_2_ limits are not exceeded and that ventilation measures are sufficient to reduce the CO_2_ levels in the room.

While our study focuses on school classrooms, these results can in principle be transferred to similar situations in closed rooms that are occupied by more than a single person, such as meeting rooms, restaurants, bars, shared offices, waiting rooms and others.

## Data Availability

The original data is available from the corresponding author upon request.

## Acknowledgments

We thank the Leibnizschule Wiesbaden for support and cooperation. We would like to thank the dean, Rainer Guss, the classes 9c and 9d, their teachers, as well as Dr. Bundschuh for permitting us to conduct the measurements and for comprehensive support of the measurements and participation in the surveys. We thank the Ministry of Education and Religious Affairs for support. We thank Dr. A. Kürten, S. Richter and T. Keber for valuable input, discussion and technical support. We thank Prof. H.-M. Seipp, Univ. Gießen, Prof. H.-J. Schmid, Univ. Paderborn, and M. Westerhoff for discussion.

## Supplemental Online Material

### 1. Time series of the uCPC measurements

**Figure S1:**
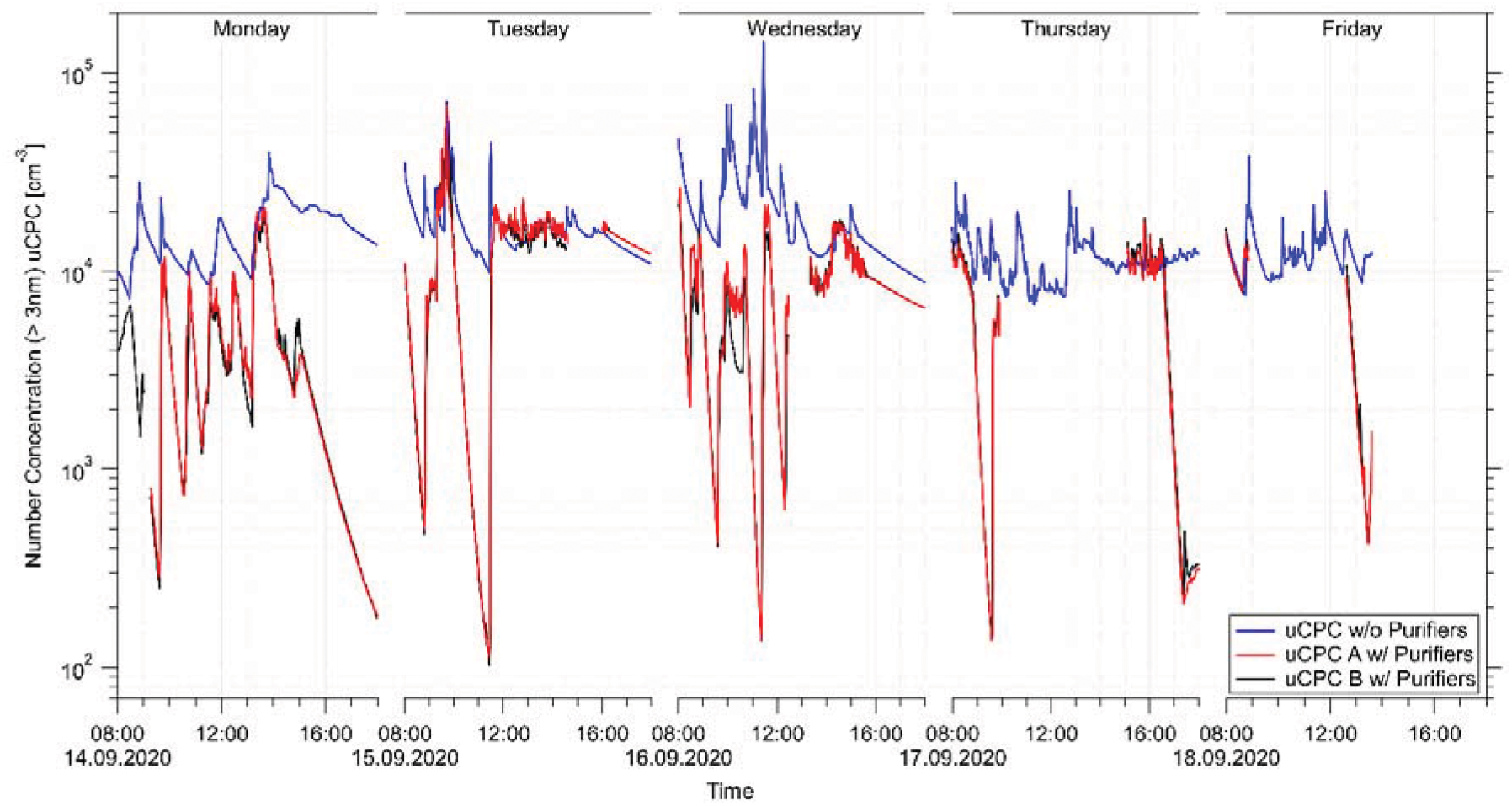
Complete time series of the uCPC measurements at Leibnizschule. The blue line shows the data from the measurements in class room B110 without operating any air purifiers. The red and black line show the data in class room B109, where on Monday, 14 Sept., 3 air purifiers were operated and this was increased to 4 air purifiers starting on Wednesday, 16 September.

### 2. Use of air purifiers and window venting for reduction of virus-containing aerosols

Here we compare different measures to reduce the concentration of potentially virus-containing aerosols in a typical class room. In addition to showing the effect of four air purifiers with HEPA filters we also show the effect of venting by opening the windows every 20 minutes for 3 minutes as it is currently recommended for schools by the German commission for indoor hygiene (IRK, 2020). While the effect of the continuous operation of the air purifiers can be specified fairly precisely if the CADR is known, the window venting depends on several parameters such as the temperature difference between outside and inside, the wind direction and wind speed, the number and size of the windows that can be opened, and the exact duration of venting. We expand the calculations shown in Figure 6 of the main text using the same general settings (class room volume 180 m^3^; total CADR of air purifiers 1026 m^3^; presence of one highly infective person, breathing with a rate of 10 liters per minute and exhaling 0.6 cm^-3^ aerosol particles of 5 µm wet size when speaking (50% of the time), and 0.06 cm^-3^ of 5 µm when just breathing (other 50%); the exhaled aerosol particles are assumed to contain 5 x 10^8^ RNA copies of the virus per milliliter of fluid; 24 non-infected persons in the room breathe as well with a rate of 10 l per minute). For the venting we assume a broad range of efficiency, in the first case realizing a reduction of 30% of the virus-containing aerosol during 3 minutes of venting, and in the second case a 99% reduction is assumed, respectively. The first case represents a situation with just low temperature difference between outside and inside, low wind speeds, or just a few windows being opened, while the second case represents a highly efficient venting when a high temperature difference is present and many large windows are opened.

Figure S2 shows the resulting time series for the concentration of virus-containing aerosols in the room, while Figure S3 shows the resulting dose that is taken up by the recipients over the time of 1 hour in the room. After 1 hour the dose of virus-RNA copies that are taken up amounts to 5.2 (closed room), 3.2 (- 30% venting), 1.5 (−99% venting), 1.5 (air purifiers), 1.3 (air purifiers and -30% venting) and 0.9 (air purifiers and -99% venting). The 30%-efficient venting reduces the inhaled dose by about 33%, while the 99% venting and the purifiers alone reduce it by 71%. The best reduction is achieved by the combined measures, totaling a reduction of the dose of 83%. Of course, it has to be considered that exchanging the room air almost completely by venting also means that the 180 m^3^ (corresponding to 229 kg) of air entering in winter time at temperatures typically below 5°C have to be warmed to 20°C which yields an energy need of more than 3.5 MJ for each venting that has to be supplied by the heating system.

In summary it can be stated that the highly efficient venting every 20 minutes alone produces about the same overall dose in one hour as the air purifiers with a CADR of 1026 m^3^/h alone. The best reduction is achieved by a combination of the measures. In this way the advantages of both safety measures are combined, yielding a continuous and constant reduction that is independent of the outside conditions from the air purifiers, as well as the reduction from the venting, which is also reducing the CO_2_ levels in the room. Additional measures, such as the highly recommended use of face masks, will reduce the risks further but are not considered here.

**Figure S2:**
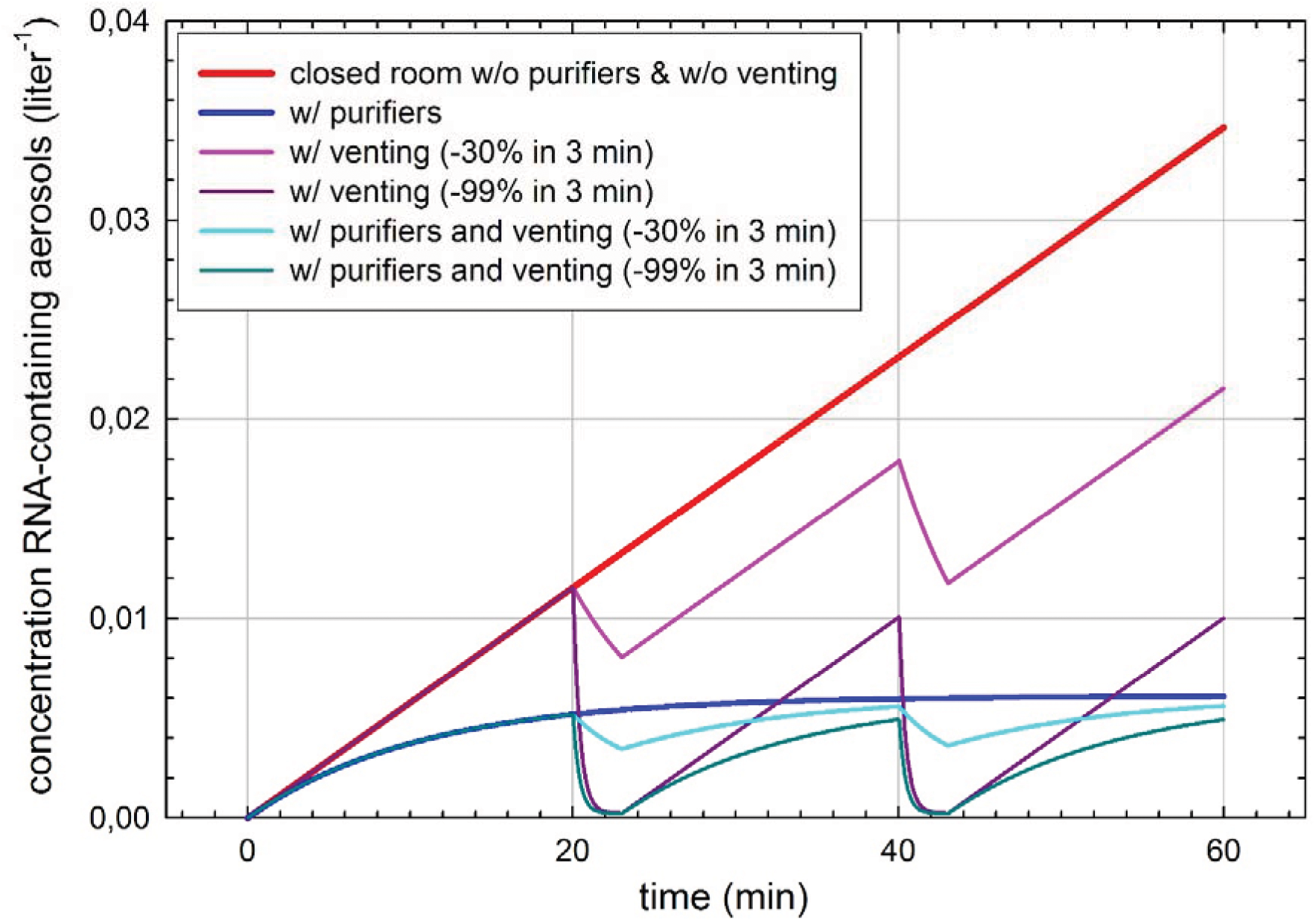
Time series of the concentration of virus-containing aerosol particles in a class room with a highly-infective person present, speaking 50% of the time. Closed room without venting or air filtration (red); closed room with continuous use of air filters with CADR of 1026 m^3^/h (blue); venting every 20 minutes for 3 minutes with low (−30%, pink) and high (−99%, purple) efficiency, and combined filtering and venting (cyan and green).

**Figure S3:**
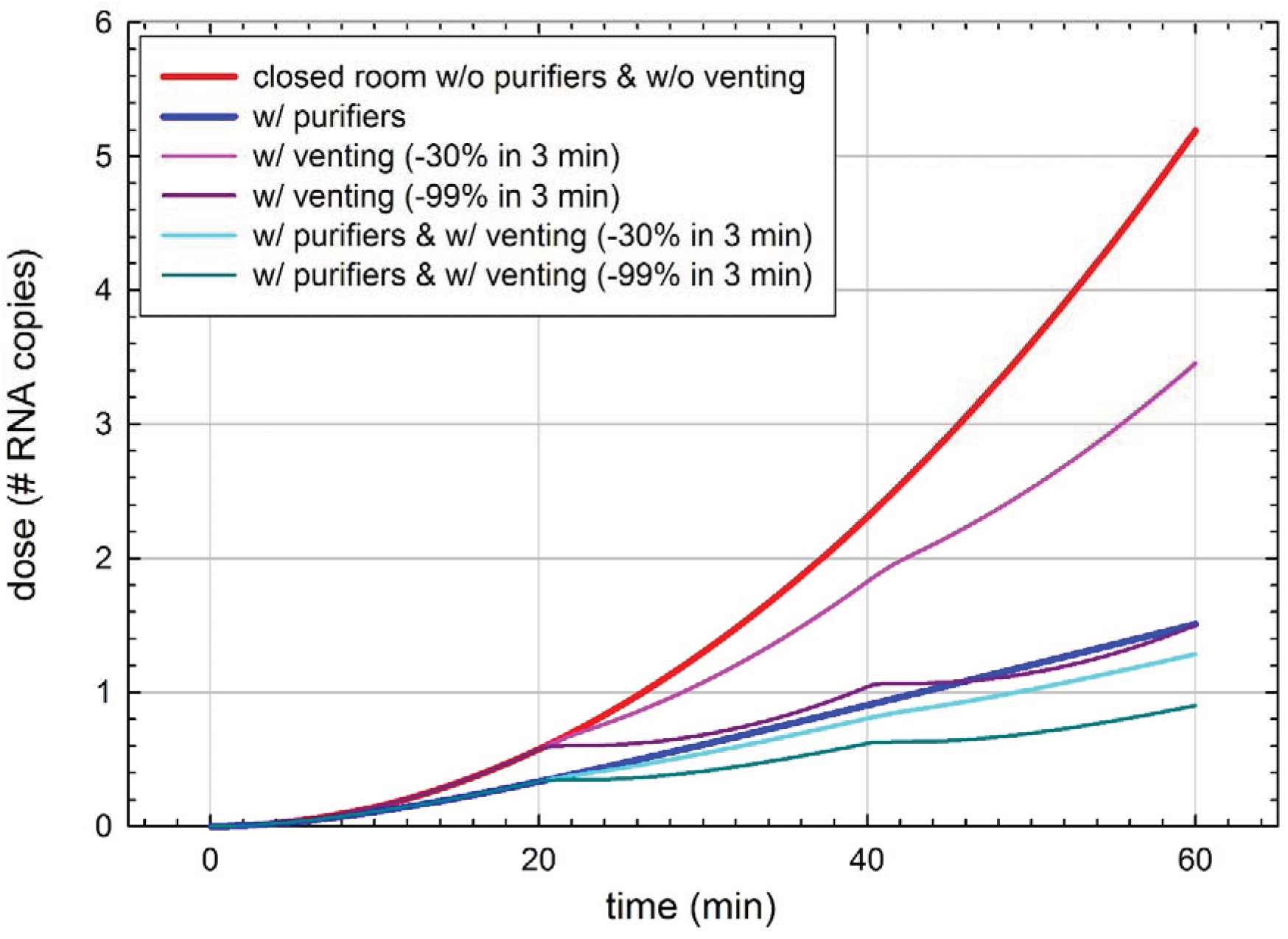
Dose of RNA copies taken up by a susceptible person in the room over 1 hour for the same conditions as in Figure S2.

### 3. Noise levels

**Figure S4:**
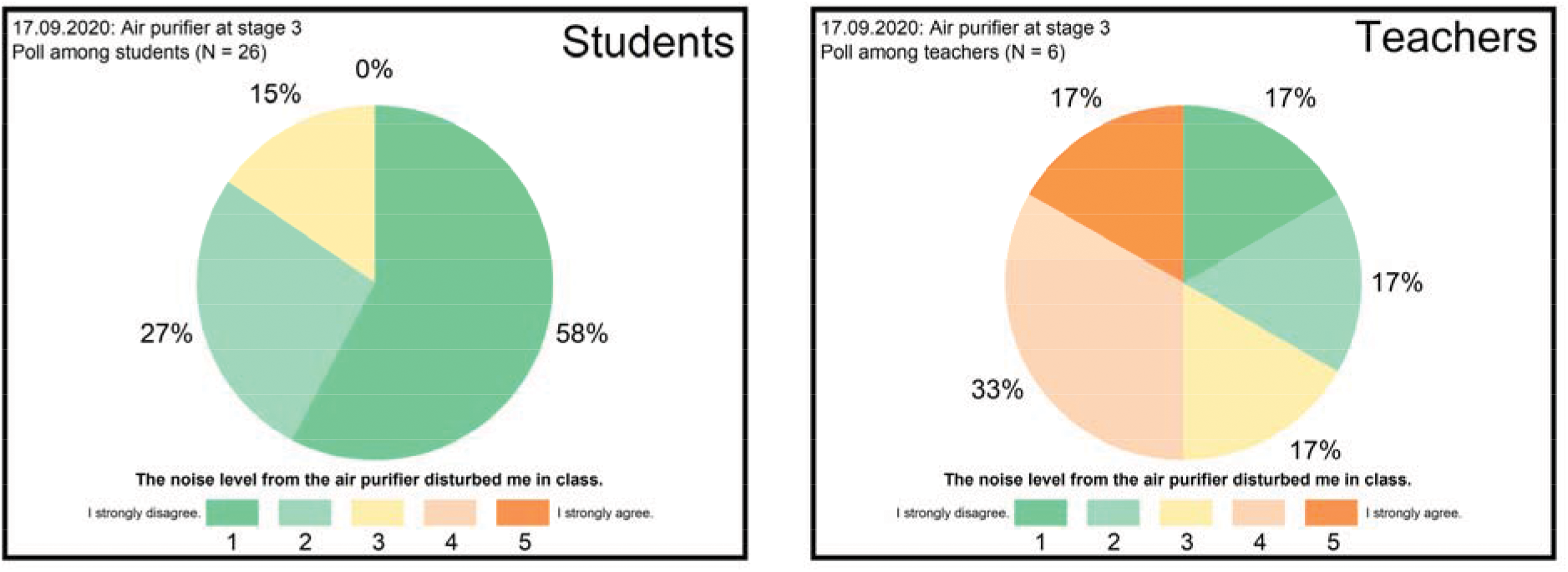
Results of the survey among students (left, n=26) and teachers (right, n=6) on disturbances by the noise levels produced by the purifiers when running four purifiers at stage 3 (total volume flow 1026 m^3^/h, air exchange rate 5.5 h^-1^).

### 4. Cleaning and Maintenance

**Figure S5:**
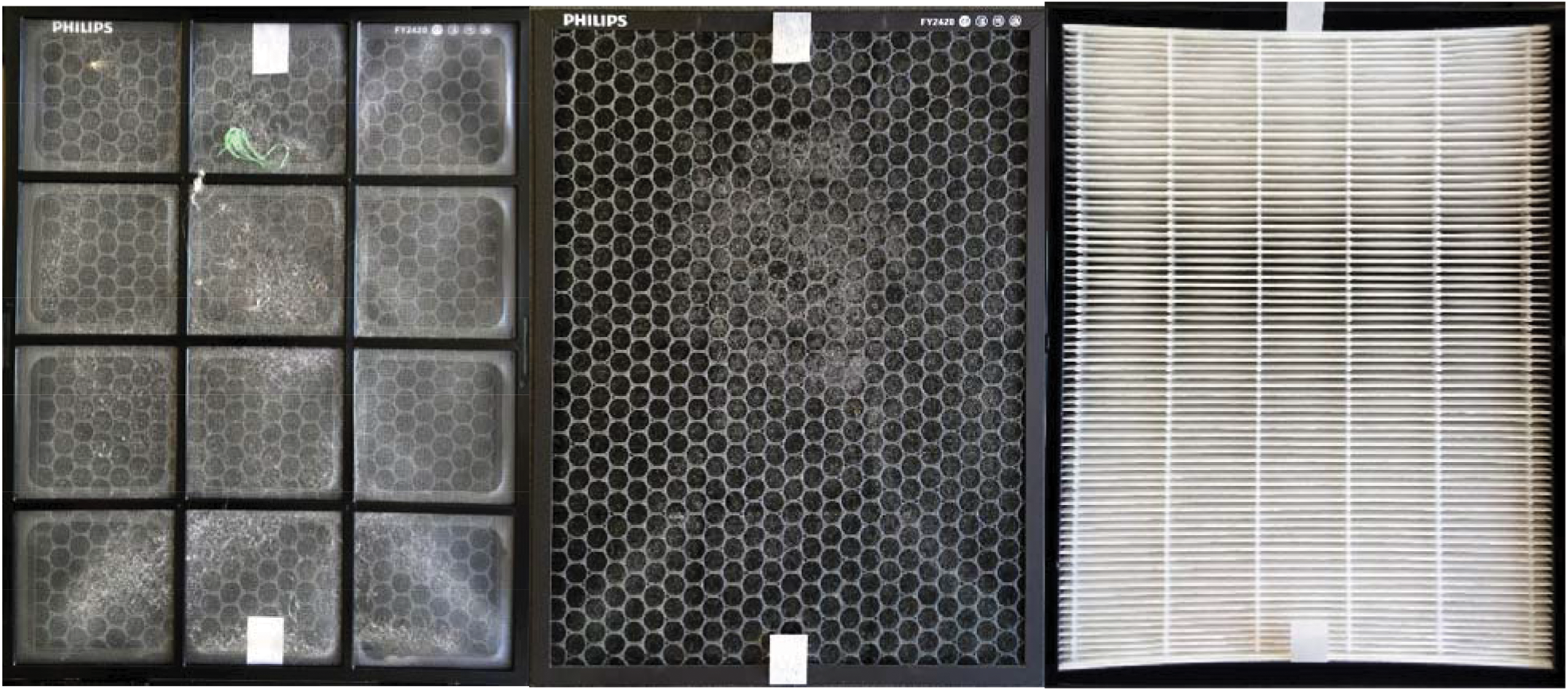
Pre-filter (left), active charcoal filter (middle) after one week of operation in the classroom. Coarse dust, hairs and fluff can be discerned. No deposits of particles could be discerned by eye on the HEPA-Filter (right). Sections that appear darker are due to the illumination.

